# Plasma proteomic associates of infection mortality in UK Biobank

**DOI:** 10.1101/2024.01.21.24301569

**Authors:** Michael Drozd, Fergus Hamilton, Chew W Cheng, Patrick J Lillie, Oliver I Brown, Natalie Chaddock, Sinisa Savic, Khalid Naseem, Mark M Iles, Ann W Morgan, Mark T Kearney, Richard M Cubbon

## Abstract

**Background:** Infectious diseases are a major cause of mortality in spite of existing public health, anti-microbial and vaccine interventions. We aimed to define plasma proteomic associates of infection mortality and then apply Mendelian randomisation (MR) to yield biomarkers that may be causally associated.

**Methods:** We used UK Biobank plasma proteomic data to associate 2,923 plasma proteins with infection mortality before 31^st^ December 2019 (240 events in 52,520 participants). Since many plasma proteins also predict non-infection mortality, we focussed on those associated with >1.5-fold risk of infection mortality in an analysis excluding survivors. Protein quantitative trait scores (pQTS) were then used to identify whether genetically predicted protein levels also associated with infection mortality. To conduct Two Sample MR, we performed a genome-wide association study (GWAS) of infection mortality using UK Biobank participants without plasma proteomic data (n=363,953 including 984 infection deaths).

**Findings:** After adjusting for clinical risk factors, 1,142 plasma proteins were associated with risk of infection mortality (false discovery rate <0.05). 259 proteins were associated with >1.5-fold increased risk of infection versus non-infection mortality. Of these, we identified genetically predicted increasing MERTK concentration was associated with increased risk of infection mortality. GWAS for infection mortality revealed no SNPs achieving genome-wide statistical significance (p<5×10^-8^). However, MR supported a causal association between increasing plasma MERTK protein and infection mortality (odds ratio 1.46 per unit; 95% CI 1.15-1.85; p=0.002).

**Interpretation:** Plasma proteomics demonstrates many proteins are associated with infection mortality. MERTK warrants exploration as a potential therapeutic target.

## Introduction

In spite of the substantial impacts achieved by public health, anti-microbial and vaccine interventions, infectious diseases remain an important cause of death across the world. For example, the Global Burden of Disease Study found that almost 20% of deaths in 2017 were sepsis-related, with marked geographical variation in the incident and fatal sepsis.(1) The risk of infection death also varies markedly within countries, with data from the United Kingdom finding that factors including advancing age, socio-economic deprivation (SED) and multimorbidity are associated with greater risk of infection than non-infection death.(2) Whilst socio-economic factors are likely to play a significant role in these disparities, biological factors are also important, for example via altered immune responses to pathogens. Indeed, genome-wide association studies (GWAS) have highlighted immune-related genes, amongst others, as being associated with risk of incident infection.(3,4) Extensive research in COVID-19 also shows an important role for host factors in relation to outcome.(5,6) In light of the risk of future pandemics, growing anti-microbial resistance (AMR), climate change, urbanisation and demographic shifts,(7) understanding the biology of host factors associated with fatal infection is an important goal. To address this, we used the recently released UK Biobank (UKB) plasma proteomics (PP) resource (also known as the UKB Pharma Proteomics Project) to define circulating factors that may represent biomarkers or therapeutic targets for infection mortality.

## Methods

### Study design and cohort

The UK Biobank is a large scale, prospective population-based study that recruited over 500,000 individuals aged 37 to 73. Recruitment occurred from 2006 to 2010, with participants visiting one of 22 assessment centres across England, Scotland, and Wales. Baseline biological measurements were collected, and individuals participated in both touchscreen and nurse-led questionnaires, as previously described.(8) Full details of the study design and conduct are available on the UK Biobank website (https://www.ukbiobank.ac.uk). All participants gave their written consent and UK Biobank received ethical approval from the NHS Research Ethics Service (11/NW/0382). Our analysis was conducted under application number 98520.

### Plasma proteomics data

The UK Biobank Pharma Proteomics Project (UKB-PPP) is a collaboration between UKB and thirteen biopharmaceutical companies that measured 2,923 unique proteins in a subset of 53,029 UKB participants. This is one of the world’s largest proteomic studies to date. Plasma proteins were measured using the antibody based Olink Explore 3072 Proximity Extension Assay technology. Detailed methodology, quality control procedures and validation are described elsewhere.(9) Protein concentrations were normalized producing NPX values (Olink’s arbitrary unit in log_2_ scale) for each protein per participant. Values below the limit of detection were classified as missing. In the present analyses we used protein measurements at baseline visit.

### Assessment of socio-demographic factors and comorbidity

Potential confounding factors considered in adjusted analyses included age, sex, ethnicity, smoking, socioeconomic status and comorbidity. These were all determined at study recruitment and our previous work has shown these are associated with infection mortality.(2) Using the same approach, we grouped ethnicity into White or Minority ethnicity (participants that self-identified as belonging to a UK Biobank defined category of Asian, Black, Chinese, Mixed or Other ethnic group). Smoking status was categorised as never, former and current. SED was measured with the Townsend score and categorised into quintiles. Obesity was determined based on the World Health Organization’s classification system based on body mass index (BMI): class 1 obesity (30.0–34.9 kg/m²), class 2 obesity (35.0– 39.9 kg/m²), and class 3 obesity (≥ 40 kg/m²). As we have previously described, medical conditions recorded during face-to-face interview with a nurse were used to classify comorbidities into disease groups including: hypertension, chronic heart disease (ischaemic heart disease and heart failure), chronic respiratory disease, diabetes, cancer (previous or current), chronic liver disease, chronic kidney disease, previous stroke or transient ischaemic attack, other neurological disease, psychiatric disease, and chronic inflammatory and autoimmune rheumatic disease.(2)

### Mortality ascertainment

Mortality information on UKB participants is received from NHS England for participants in England & Wales and from the NHS Central Register a component of the National Records of Scotland, for those in Scotland. In the present analysis, we focused on deaths due to infection prior to the COVID-19 era and therefore censored follow-up until 31st December 2019 to ensure this was before the first reported case of Coronavirus 2019 in the UK.(10) This approach was chosen to avoid the risk of biasing our data by including a substantial proportion of deaths attributed to one pandemic pathogen. Infection deaths were classified based on the ICD-10 coding system as we have previously described.(2)

### Plasma protein association with cause-specific mortality

We estimated adjusted cause-specific mortality incidence rate ratios (IRRs) for specific proteins using Poisson regression models with exposure time modelled. Fully adjusted models included: age, sex, SED, smoking, obesity, hypertension, chronic heart disease (ischaemic heart disease and heart failure), chronic respiratory disease, diabetes, cancer (previous or current), chronic liver disease, chronic kidney disease, previous stroke or transient ischaemic attack, other neurological disease, psychiatric disease, and chronic inflammatory and autoimmune rheumatic disease. Age was modelled using restricted cubic splines – five knots for infection mortality analyses and four knots for non-infection mortality analyses. This approach was selected as it provided the best fit as assessed by the Akaike information and Bayesian criteria (comparing various models including categorical, linear, cubic splines, and first and second-degree fractional polynomials). To identify proteins more specifically associated with infection mortality than non-infection mortality, we repeated the fully adjusted models after excluding all participants that survived to the censorship date (31st December 2019). In this case, IRR>1 indicates that the protein is more specifically associated with increased risk of infection mortality than non-infection mortality (and *vice versa* for IRR<1). In all analyses, multiple testing was accounted for using Benjamini-Hochberg FDR adjusted p values, defining statistical significance as FDR-adjusted p<0.05.

### Functional Enrichment of plasma proteins associated with infection mortality

Functional enrichment analysis of hits was performed using g:Profiler (version e110_eg57_p18_4b54a898; https://biit.cs.ut.ee/gprofiler/gost)(11) with Benjamini-Hochberg FDR adjustment of p values for identified pathways. Functional profiling was performed on statistically significant proteins with IRR >1.5 in the infection mortality specificity analysis (i.e. those associated with greater risk of infection mortality compared to non-infection mortality). A custom background of 2,923 genes representing the proteins measured in the UKB-PPP were set as the statistical domain.

### Plasma protein polygenic scores

We utilised validated Olink plasma protein quantitative trait scores (pQTS) developed by Xu *et al* (www.omicspred.org).(12) These genetic scores were derived from a subset of the INTERVAL cohort using Bayesian Ridge Regression with validation on NSPHS and ORCADE cohorts. Genetic scores were available for 46 of the 259 proteins that we identified as being most specifically associated with infection mortality. We applied these scores to each UKB participant of white British ancestry who was not in the plasma proteomics cohort (to avoid bias due to sample overlap) using the sum of weighted alleles. We excluded participants with sex discrepancies between reported sex and genetic sex, sex chromosome aneuploidy, outliers for heterozygosity or missing rate and individuals with a missing genotype rate >10%. These polygenic scores were then included in Poisson regression models (as described earlier) for infection and non-infection mortality adjusted for age, sex and the first ten genetic principal components to account for population stratification.

### Mendelian randomisation

Mendelian randomisation is an approach that uses the random allocation of alleles at conception to generate estimates of the causal effects on exposures on an outcome.(13) It has three core assumptions which are required for any instrumental variable approach. Firstly, that the genetic variation is associated with the exposure (relevance); secondly, that there are no causes of the genetic variation that have an effect on the outcome except through the exposures (exchangeability); and thirdly, that the genetic variation acts through the exposure only (exclusion restriction). We applied two-sample MR analyses to estimate the predicted causal effects of levels of plasma proteins (exposure) and infection mortality (outcome) using the TwoSampleMR 0.5.8 package (14) Instruments for plasma protein levels were extracted from a recent GWAS of MERTK levels performed in the UK Biobank proteomic cohort.(9) To reduce the risk of pleiotropy, these were pruned to *cis* genetic instruments by selecting genome wide significant (p<5×10^-8^) uncorrelated SNPs (LD in the European subset of the 1000 Genomes Project R^2^ <0.1) within 500Kb of the transcript boundaries of the respective gene encoding the protein.(15)

### Infection mortality genome-wide association study (GWAS)

To identify genetic estimates in our outcome sample, we performed a case-control GWAS with infection mortality as the outcome to generate summary statistics for downstream Mendelian randomisation (MR) analyses. We performed this in the sample excluding those with measured proteomic data (n=363,953), as sample overlap in MR can lead to biased estimates, but a sensitivity analysis also used the full cohort (n=407,264). The degree of bias in the latter approach would be expected to be small in this case as sample overlap is not complete.(16) We included participants of White British ancestry (UKB Field 22006) and conducted GWAS using regenie v3.1.1.(17) Quality control and full methodological details are available in **Supplementary Data 1**.

### MR approach and sensitivity analyses

Effect estimates from the exposure and outcome GWAS were identified and effect alleles harmonised. MR estimates for each SNP were generated by the Wald ratio and meta-analysed by the inverse-variance weighted (IVW) method in our primary analysis.(18). We additionally performed a number of sensitivity analyses: a) relaxing the LD threshold to 0.2, to allow more instruments but potentially allowing correlated SNPs, tightening the LD threshold to 0.01, to ensure SNPs were not correlated, c) performing meta-analysis including a correlation matrix to allow for correlated SNPs, d) including in the outcome GWAS participants who had measured levels of proteomic data, e) reporting other meta-analytic approaches (e.g. MR-Egger and Weighted Median), and d) performing Radial MR to identify outliers. Analyses were performed using the TwoSampleMR, Radial MR, and MendelianRandomization packages.(14,18–20)

### Statistical analyses

Continuous variables are presented as mean (SD) or median (IQR) if non-normally distributed. These were visually assessed using histograms and Q-Q plots. Categorical variables are presented as number (%). All statistical tests were two-sided and statistical significance was defined as p<0·05. Analyses were performed in R (v4.1.1) with Poisson models implemented using the glm function.

## Results

The UKB cohort study includes baseline PP data for 53,029 participants, of whom we excluded 509 (0.96%) due to missing data or absent long-term follow-up data. Within this subgroup, 240 infection deaths and 3,551 non-infection deaths occurred before 31^st^ December 2019, during 558,616 person years of follow-up (median 10.8 [IQR 10.2-11.6] years per participant). The most common fatal infections involved the lower respiratory tract (136 events [57.6%]), gastrointestinal tract (39 events [16.5%]), and genitourinary tract (14 events [5.9%]). We defined the association between increasing concentrations of 2,923 distinct plasma proteins and infection mortality, in models including baseline socio-demographic factors and comorbidities known to be associated with infection mortality. After accounting for multiple testing, 1,142 proteins were associated with the risk of infection mortality, of which 1,110 were with higher risk and 32 with lower risk (**Figure 1A** and **Supplemental Data 2A**). A complementary analysis applied identical models to define associations of the 2,923 proteins with non-infection mortality. After accounting for multiple testing, 1,334 proteins were associated with the risk of non-infection mortality, of which 1,196 were with higher risk and 138 with lower risk (**Figure 1B** and **Supplemental Data 2A**). Hence, it is likely that a substantial proportion of the proteins associated with infection mortality are non-specific associates of mortality *per se*. As we aimed to focus on those proteins most specifically associated with infection mortality, we repeated our analysis of proteins associated with infection mortality after excluding all subjects who survived during the observation period. Therefore, any protein demonstrating an IRR significantly above one would show greater specificity for infection mortality and any demonstrating an IRR significantly below one would show greater specificity for non-infection mortality. Applying a threshold of IRR>1.5 in this analysis allowed us to focus on proteins with greater specificity for infection mortality and after accounting for multiple testing 259 proteins reached statistical significance (**Figure 2** and **Supplemental Data 2B**). To explore the biological themes linking these proteins, we conducted GO analysis of these 259 proteins, which revealed ‘Signalling Receptor Activity’ (GO:0038023) and ‘Molecular Transducer Activity’ (GO:0060089) as the most statistically significant terms (FDR-adjusted p=2.4×10^-9^); other enriched GO terms are listed in **Supplemental Data 2C**. These data suggest that the abundance of many plasma proteins is associated with risk of infection mortality, although with only broad biological themes linking these in enrichment analysis.

**Figure 1:**
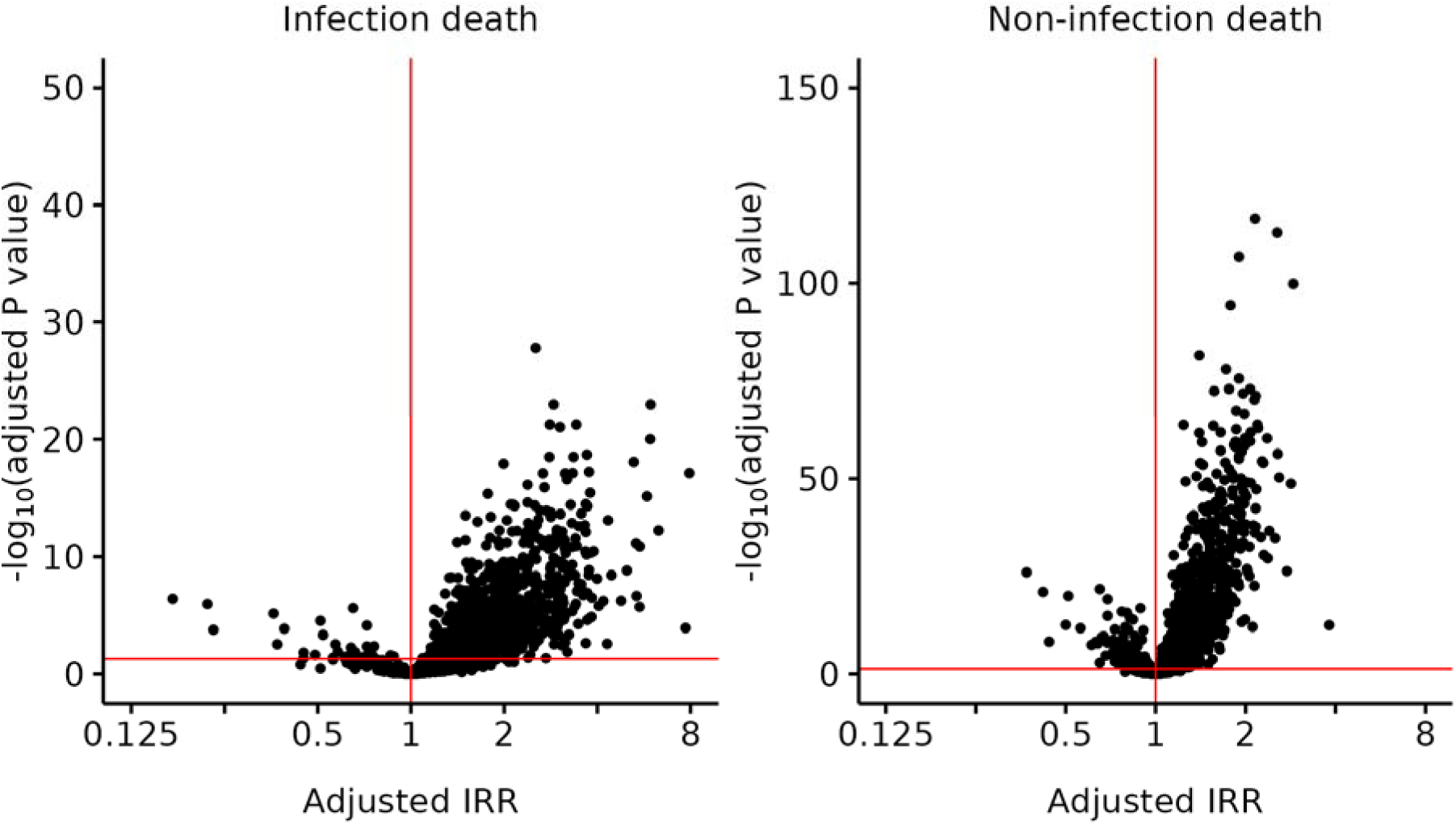
Plasma proteomic associates of infection and non-infection mortality. Volcano plots illustrating association between concentrations of 2,933 plasma proteins and infection (left) or non-infection (right) mortality. Each dot represents one protein, with points above the red horizontal lines achieving statistical association with mortality after accounting for multiple testing (Benjamini-Hochberg FDR-adjusted p<0.05). Incidence rate ratios (IRR) above 1 indicate increasing concentration of a protein are associated with increased mortality risk and *vice versa*.

**Figure 2:**
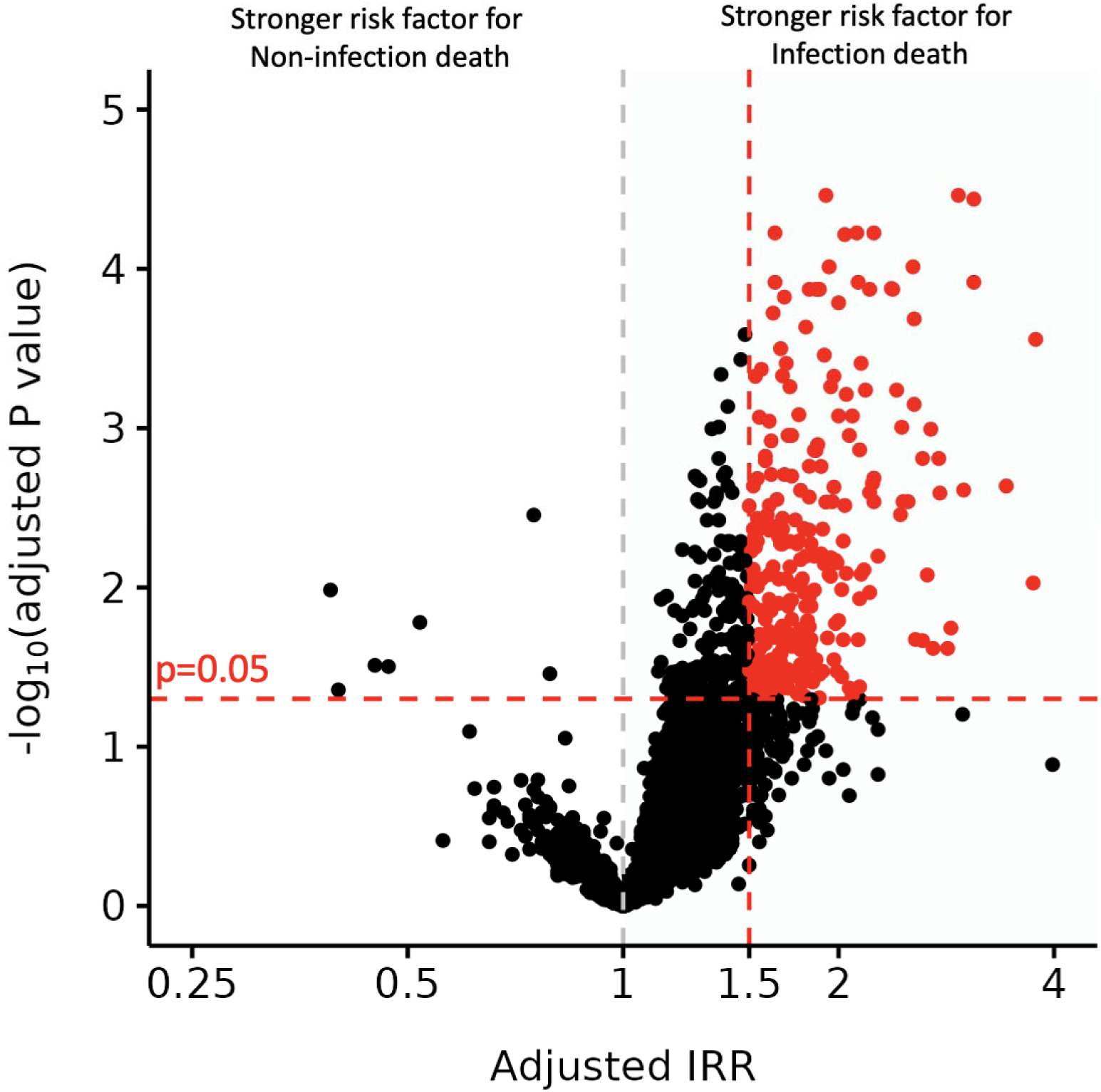
Plasma proteins with greater specificity for infection mortality. Volcano plots illustrating association between concentrations of 2,933 plasma proteins and infection mortality after excluding survivors. Each dot represents one protein, with points above the red horizontal line achieving statistical association with infection mortality after accounting for multiple testing (Benjamini-Hochberg FDR-adjusted p<0.05). Incidence rate ratios (IRR) above 1 indicate higher concentrations of proteins being mor specifically associated with increased mortality risk and *vice versa*. Dots in red, with IRR>1.5, are highlighted as proteins having greater specificity for infection mortality.

To corroborate these observations, we used validated pQTS from the OMICSPRED study as a genetic proxy for circulating protein concentrations in all UKB participants without plasma proteomic data.(12) This gave an independent cohort of 363,953 UKB participants with white British ancestry, in whom in whom 984 infection deaths occurred. The OMICSPRED study was able to generate validated pQTS for 46 of the 259 protein hits described above. For each pQTS, we defined its association with infection mortality, in models including baseline age, sex, and the first ten genetic principal components (**Supplemental Data 2D**). A parallel analysis was conducted for non-infection mortality using an otherwise identical approach (**Supplemental Data 2D**). Only the pQTS for MERTK was significantly associated with infection mortality (IRR 1.38; 95% CI 1.08-1.76; FDR-adjusted p=0.010); this was not associated with non-infection mortality (1.02; 95% CI 0.96-1.07; FDR-adjusted p=0.581).

To infer whether the association between plasma MERTK protein and infection mortality may be causal, we performed MR. Since there is no published GWAS of infection mortality, we first conducted this in the UKB white British population without PP data to allow downstream two-sample MR. This analysis included 984 cases (i.e. infection mortality) and 362,969 controls. Within 15,946,334 genetic variants assessed, we found none that reached a genome-wide level of statistical significance (**Figure 3**, **Supplemental Figure 1** and **Supplemental Data 2E**). However, 3 SNPs exceeded the threshold for genome-wide suggestive significance (p<1×10^-6^): rs76098597, rs547524815 and rs6694475. Of these, the nearest genes for the former two are non-coding, although rs6694475 is in the locus of *Usp48*, a gene that has been implicated in pathogen responses.(21,22)

**Figure 3:**
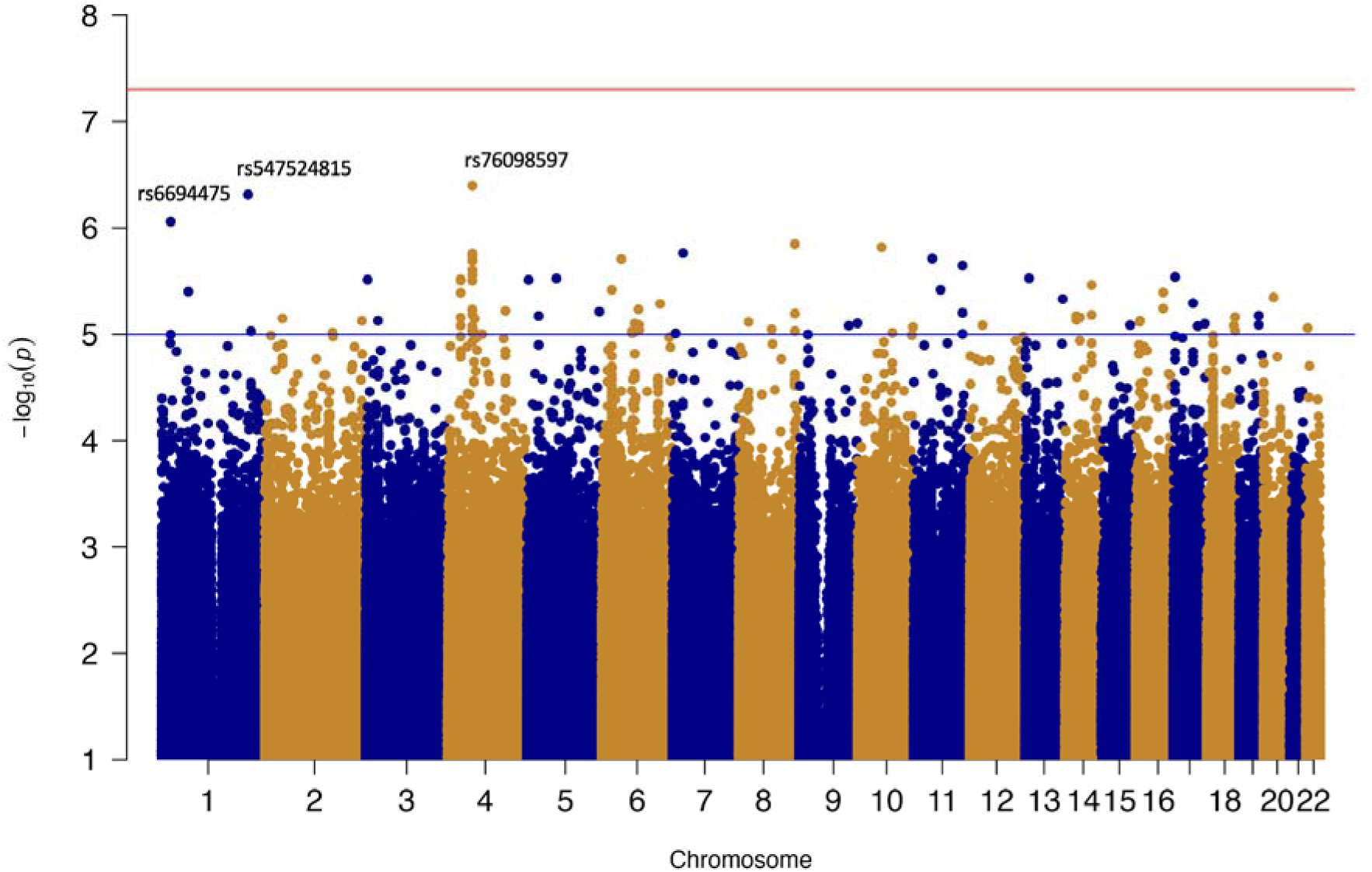
Genome-wide association study for infection mortality. Manhattan plot illustrating association between 15,946,334 single nucleotide polymorphisms (SNPs, represented as grey and blue dots) and infection mortality. The red line denotes the threshold for genome wide statistical significance (p<5×10^-8^), which no SNPs achieve. The blue line denotes the threshold for suggestive significance (p<1×10^-5^).

We then conducted two-sample MR using summary statistics from our infection mortality GWAS, together with instruments derived from UK Biobank.(9) Our primary instrument included 23 uncorrelated (R^2^ >0.1) SNPs *cis* to MERTK. The minimum F-statistic was 36.1. All SNPs are reported in **Supplemental Table 2F**. IVW MR estimates suggested a predicted causal effect of increased plasma MERTK concentration on infection mortality (Odds ratio (OR) per standard deviation (SD) increase 1.46; 95% CI 1.15-1.85; p=0.002; **Figure 4**). Sensitivity analyses using different meta-analytic approaches yielded similar estimates (**Figure 4**). We re-ran analyses using different definitions of our instrument, and accounting for SNP correlation using a correlation matrix; these results are reported in **Supplemental Table 2G**. MR results from analyses that were more conservative in SNP selection (R^2^>0.01), or when accounting for SNP correlation effect estimates using a correlation matrix, yielded similar estimates, but with wider confidence intervals. MR results were very similar in a sensitivity analysis using the GWAS that included sample overlap (**Supplemental Table 2H**), likely reflecting strong instruments (median F-statistic 69.1), and limited sample overlap. Finally, Radial MR did not identify any SNP outliers (**Supplemental Figure 2**). Collectively, these data support a potentially causal association between higher plasma MERTK protein and increased odds of infection mortality.

**Figure 4:**
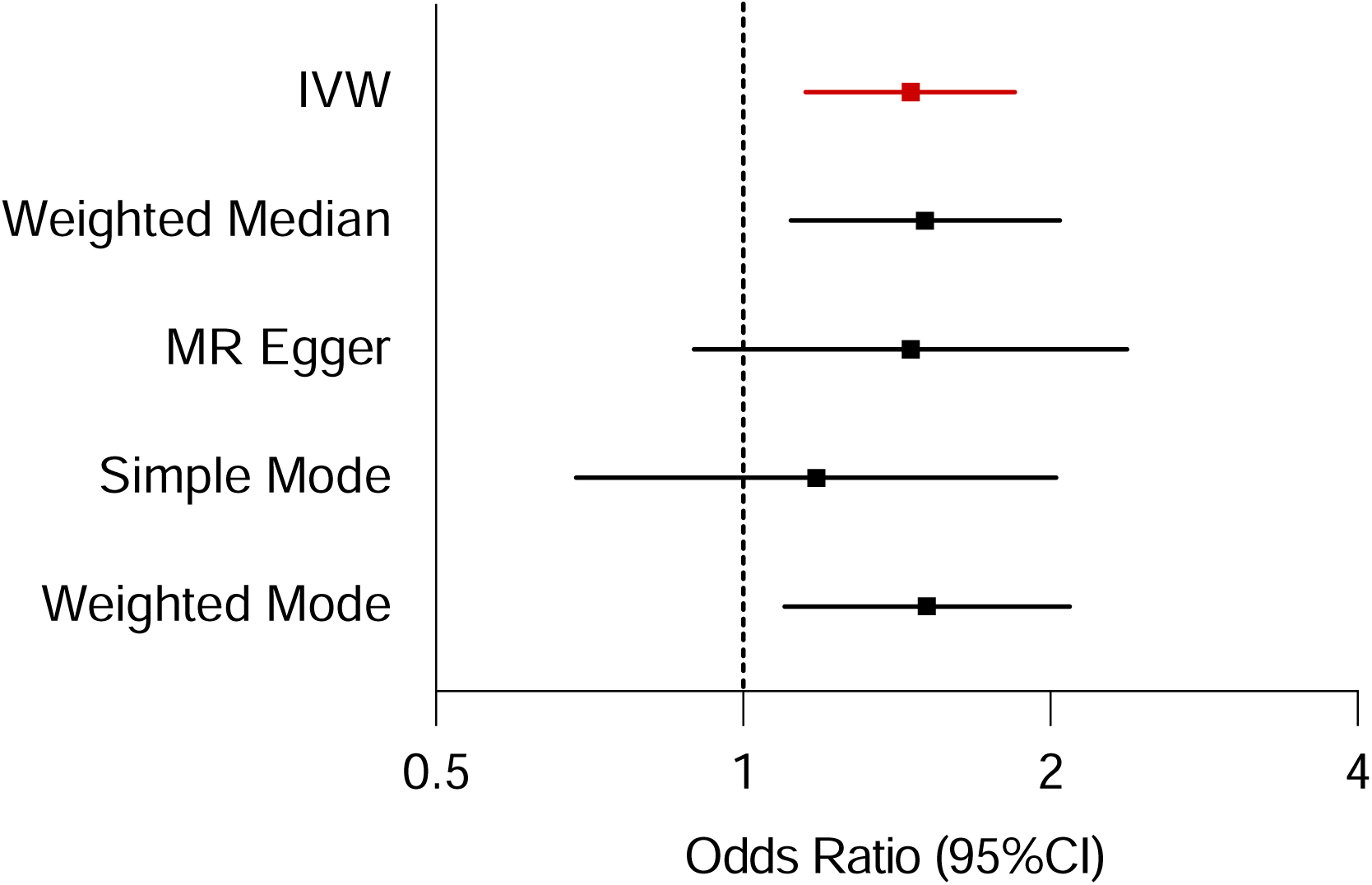
Mendelian randomisation of plasma MERTK association with infection mortality. Forest plot illustrating MR estimates of plasma MERTK protein association with infection mortality. Primary instrument included 23 uncorrelated (R2 >0.1) SNPs cis to MERTK. Bar represents 95% CI.

Finally, we divided the UKB-PP cohort into quartiles of MERTK to understand the characteristics of participants with higher plasma MERTK concentrations (**Table 1**). This revealed that higher MERTK was associated with older age, male sex, and multiple chronic illnesses, especially cardio-metabolic disorders. Whilst statistically significant, plasma highly sensitive C-reactive protein (hsCRP), a biomarker of low-grade inflammation, was only modestly increased with rising MERTK. There were also modest, but statistically significant, associations between plasma MERTK and circulating leukocyte subsets and platelet counts.

**Table 1:** Participant characteristics in MERTK quartiles at study recruitment

|  | **Q1**<br>**(n=12899)** | **Q2**<br>**(n=12899)** | **Q3**<br>**(n=12899)** | **Q4**<br>**(n=12899)** |
| --- | --- | --- | --- | --- |
| Age, years | 53.9 (0.1) | 56.2 (0.1) | 57.7 (0.1) | 59.5 (0.1) |
| Male sex | 4345(18.3%) | 5708(24.0%) | 6470(27.3%) | 7212(30.4%) |
| Obesity |  |  |  |  |
| Class 1 | 1906 (14.8%) | 2193 (17%) | 2394 (18.6%) | 2555 (19.8%) |
| Class 2 | 549 (4.3%) | 593 (4.6%) | 664 (5.1%) | 784 (6.1%) |
| Class 3 | 206 (1.6%) | 240 (1.9%) | 256 (2%) | 343 (2.7%) |
| Hypertension | 2661 (20.6%) | 3135 (24.3%) | 3773 (29.3%) | 4734 (36.7%) |
| Chronic heart disease | 401 (3.1%) | 563 (4.4%) | 698 (5.4%) | 1144 (8.9%) |
| Respiratory disease | 1738 (13.5%) | 1683 (13%) | 1660 (12.9%) | 1762 (13.7%) |
| Diabetes Mellitus | 530 (4.1%) | 580 (4.5%) | 702 (5.4%) | 999 (7.7%) |
| Cancer | 1005 (7.8%) | 1056 (8.2%) | 1080 (8.4%) | 1214 (9.4%) |
| Liver disease | 23 (0.2%) | 22 (0.2%) | 15 (0.1%) | 61 (0.5%) |
| Chronic kidney disease | 19 (0.1%) | 26 (0.2%) | 46 (0.4%) | 102 (0.8%) |
| Stroke | 166 (1.3%) | 202 (1.6%) | 258 (2%) | 407 (3.2%) |
| Neurological disease | 141 (1.1%) | 201 (1.6%) | 245 (1.9%) | 355 (2.8%) |
| Psychiatric disease | 798 (6.2%) | 753 (5.8%) | 811 (6.3%) | 846 (6.6%) |
| Rheumatological disease | 280 (2.2%) | 335 (2.6%) | 346 (2.7%) | 496 (3.8%) |
| Leukocytes, 10<sup>9</sup> cells/L | 6.6 (5.6-7.9) | 6.6 (5.6-7.8) | 6.7 (5.6-7.8) | 6.7 (5.7-7.9) |
| Neutrophils, 10<sup>9</sup> cells/L | 4.0 (3.2-5.0) | 4.0 (3.2-5.0) | 4.0 (3.3-4.9) | 4.1 (3.3-5.1) |
| Lymphocytes, 10<sup>9</sup> cells/L | 1.9 (1.5-2.3) | 1.9 (1.5-2.3) | 1.9 (1.5-2.3) | 1.8 (1.5-2.3) |
| Monocytes, 10<sup>9</sup> cells/L | 0.43 (0.34-0.54) | 0.45 (0.36-0.56) | 0.46 (0.37-0.58) | 0.48 (0.38-0.60) |
| Platelets, 10<sup>9</sup> cells/L | 254 (219-293) | 250 (215-289) | 246 (212-285) | 241 (207-281) |
| C-reactive protein, mg/L | 1.26 (0.61-2.76) | 1.31(0.66-2.75) | 1.35 (0.68-2.78) | 1.45 (0.72-2.96) |

Data presented as median (IQR) or number (%).

## Discussion

To our knowledge, we present the most comprehensive analysis of plasma proteomics in relation to infection mortality. This reveals many circulating proteins associated with the risk of infection mortality, a small subset of which demonstrate stronger association with risk of infection than non-infection mortality. These could represent promising biomarkers and avenues to explore the pathophysiology of infection mortality. Focussing within this subset, we sought complementary evidence for association with infection mortality and found that a polygenic score for plasma MERTK also demonstrated concordant association with infection mortality. To extend this, we conducted MR analyses, which supported the likelihood of a causal association between plasma MERTK and risk of infection mortality. Whilst the biology underpinning the association needs further evaluation, this raises the possibility that novel therapies could mitigate a predisposition for fatal infection in people with elevated plasma MERTK.

The large proportion of circulating proteins associated with infection mortality probably attests not only to the statistical power of the UK Biobank resource, but also to the complex factors predisposing to infection mortality. A broad range of socio-demographic factors and non-communicable diseases are known to be associated with increased risk of infection (and non-infection) mortality.(2) Hence, many plasma proteins are likely to be indirectly associated with infection mortality via proximate associations with factors such as ageing and comorbidity. Indeed, when we sought proteins more strongly associated with infection than non-infection mortality, this substantially reduced the number of hits. Analysis of common biological themes in this subset found ‘signalling receptor activity’ as the most significant GO term. Specific signalling cascades were not highlighted, but this may reflect the intrinsic bias in assessing plasma proteins, in spite of our analytical approach accounting for this limited search space. Notably, the GWAS we conducted as a prelude to MR does not suffer from this limited search space, although this identified no hits achieving GWAS significance. However, it has been noted that disease-specific mortality GWAS consistently yield many fewer hits than GWAS for disease onset at a comparable sample size.(23) We believe that this is the first ever published GWAS of infection mortality that has not focussed on specific pathogens. Whilst, genome-wide significant hits were not generated, genome-wide suggestive significance (p<1×10^-6^) was observed for rs6694475; this is in the locus of Usp48, a gene that has been implicated in pathogen responses.(21,22) Notably, this includes regulation of type I interferon signalling in antiviral responses.(22)

Our major finding is the potentially causal association between increased plasma MERTK and infection mortality, which was identified in both observational and genetic analyses. We used *cis*-MR to limit pleiotropy, which is recognised as the best MR approach to identify potentially causally relevant proteins and/or drug targets.(24) This protein is encoded by the *Mertk* gene, which belongs to the TAM (Tyro3, Axl, Mertk) family of receptor tyrosine kinases, which collectively have roles in dampening the innate immune response, clearance of apoptotic cells, platelet function and regulation of vascular biology.(25,26) *Mertk* is most abundantly expressed in monocytes and macrophages, although is detected in many other lineages. Activation of MERTK signalling occurs upon binding of GAS6 (Growth arrest specific-6) or PROS1 (Protein S) ligands, both of which undergo vitamin K-dependent carboxylation which promotes calcium-dependent binding to phosphatidyl-serine, a membrane lipid enriched on apoptotic cells and activated platelets. Interestingly, increasing plasma GAS6, but not PROS1, was also associated with increased risk of infection mortality (**Supplemental Data 2A**); however, neither was in the group of 259 plasma proteins with greatest specificity for infection mortality (**Supplemental Data 2B**). The potential role of these ligands, especially GAS6, in the association we have demonstrated between MERTK and infection mortality requires further exploration.

Whilst signalling by TAM family kinases can follow typical cascades (e.g. PI3K/Akt), they are unusual in their interactions with type I interferon receptors to induce JAK/STAT signalling, which may be relevant to our GWAS signal.(25,26) This interaction of activated TAM receptors with type I interferon receptors is thought to allow a transition from pro-inflammatory to anti-inflammatory interferon signalling required during resolution of the innate immune response. Interestingly, our data show no association between plasma MERTK and plasma hsCRP nor circulating leukocyte subsets, perhaps reflecting that that these are crude biomarkers of the overall inflammatory milieu. Notably, our data are likely to reflect cleaved MERTK free in plasma, and this process is induced by the ADAM17 protease, for example in response to pro-inflammatory stimuli including lipopolysaccharide (LPS) and poly I:C.(27) Importantly, inhibition of ADAM17 suppresses LPS-induced lung inflammation and augments lung MERTK phosphorylation in mice.(28) Hence increased circulating MERTK could indicate a pro-inflammatory state and diminished capacity immune dampening via MERTK, but further experimental evidence is needed to corroborate this hypothesis. However, the broader link between *Mertk* and infection has been made in a variety of murine models and human observational studies, giving further credence to our finding.(29–35) This supports a focus on MERTK and its partners in a search for therapeutics to reduce risk of fatal infection, in addition to potential roles as biomarkers.

It is also important to discuss the limitations of our study. First, the UK Biobank cohort is not nationally representative (with lower than expected levels of SED, minority ethnic groups and some chronic diseases), which may influence the strength of associations we have observed.(36) Moreover, caution should be applied when extrapolating the data to other populations, especially in relation to our genetic analyses, which focussed on the predominant ‘White British’ ancestral group to improve statistical power by reducing variability. Second, our exclusion of infection deaths after the onset of the COVID-19 pandemic means that our findings are not relevant to SARS-CoV-2 infection. This was a deliberate decision, given the many excellent studies focussing specifically on this pathogen, and the risk of biasing our data by including a substantial proportion of deaths attributed to one pandemic pathogen. Given the absence of data on non-fatal infection episodes in UK Biobank, we also cannot comment on the association of hits with predisposition to infection episodes *per se*, which may help to understand mechanisms of association with fatal infection. Finally, the use of Olink proteomic data in UK Biobank means that we cannot comment on defined plasma protein concentrations since these are quantified in terms of normalised protein expression. Whilst this is not problematic when demonstrating plasma protein associations with infection mortality, translating our findings to clinically useful biomarkers will require additional studies defining absolute protein concentrations to ascertain risk thresholds.

To conclude, we present the most comprehensive analysis of the plasma proteomics and genetics of infection mortality. After accounting for known clinical associates of infection mortality, we implicate many circulating proteins as molecular markers of risk which may help to define the underlying biology. In particular we causally implicate increased plasma MERTK and exploration of this may represent a promising approach to developing targeted therapies to reduce fatal infection in high risk populations.

## Data Availability

The UK Biobank resource is open to all bona fide researchers. The GWAS summary statistics generated will be deposited to the GWAS Catalog (https://www.ebi.ac.uk/gwas/) once the final per reviewed manuscript is published.

## Acknowledgements

This research has been conducted using the UK Biobank Resource under application 98520. This work uses data provided by patients and collected by the NHS as part of their care and support; Copyright © (2022), NHS Digital; Re-used with the permission of UK Biobank. All rights reserved. This research used data assets made available by National Safe Haven as part of the Data and Connectivity National Core Study, led by Health Data Research UK in partnership with the Office for National Statistics and funded by UK Research and Innovation (research which commenced between 1st October 2020 – 31st March 2021 grant ref MC_PC_20029; 1st April 2021-30th September 2022 grant ref MC_PC_20058). AWM, MI, MTK and RMC receive salary support from National Institute for Health and Care Research (NIHR) Leeds Biomedical Research Centre and AWM is an NIHR Senior Investigator. The views expressed are those of the author(s) and not necessarily those of the NHS, the NIHR or the Department of Health and Social Care. SS is supported by the Kennedy Trust. NC is funded by the Medical Research Council Discovery Medicines North Doctoral Training Programme. CC is supported by British Heart Foundation Mautner Career Development Fellowship.

## Transparency Statement

RMC affirms that the manuscript is an honest, accurate, and transparent account of the study being reported; no important aspects of the study have been omitted; there are no discrepancies from the study as originally planned.

## Data sharing Statement

The UK Biobank resource is open to all *bona fide* researchers. The GWAS summary statistics generated will be deposited to the GWAS Catalog (https://www.ebi.ac.uk/gwas/).

## Authorship Statement

Guarantors – MD and RMC; Study conception – MD and RMC; Data analysis – MD, CC and FH; Manuscript drafting – MD and RMC; Critical revision of manuscript: PJL, OIB, NC, SS, KN, MI, AWM, MTK. All authors had access to data and MD and RMC verified the data. The corresponding authors attest that all listed authors meet authorship criteria and that no others meeting the criteria have been omitted.

## Supplemental material summary

**Supplementary Data 1:** Genome-wide association study extended methods and quality control data (.docx)

**Supplemental Figure 1:** Q-Q plot illustrating distribution of observed versus expected p values in genome-wide association study of infection mortality.

**Supplemental Figure 2:** Radial MR plot which shows the MR effect estimate for each SNP on the Y-axis and the weight attached to it on the X-axis. This allows for visualisation of all SNP MR effect estimates, and the identification of outliers which are outside the range of the plot. In this plot we focus on the IVW estimate for the effect of MERTK on infection mortality.

**Supplemental Data 2:** Results of proteomic and genome-wide association studies in tabular format (.xlsx). **A**) Association between increasing concentrations of 2,923 plasma proteins and infection mortality or non-infection mortality; **B**) Association between increasing concentrations of 2,923 plasma proteins and specificity for infection mortality; **C**) Gene Ontology analysis of 259 plasma proteins demonstrating greater specificity for infection mortality (IRR>1.5 and FDR adjusted p<0.05 in Supplemental Data 2C; **D**) Association between 46 plasma protein quantitative trait scores and infection mortality; **E**) Summary data for 200 most statistically significant single nucleotide polymorphisms in genome-wide association study for infection mortality (with all summary statistics deposited at https://www.ebi.ac.uk/gwas/). **F)** Included SNPs for *cis-*MERTK exposure with different R^2^ thresholds; **G)** Mendelian randomisation results of MERTK exposure with infection death outcome GWAS summary statistics in UKB individuals without plasma proteomic data (no overlap); **H)** Mendelian randomisation results of MERTK exposure with infection death outcome GWAS summary statistics in all UKB individuals

