## Supplemental Data 1 for "Plasma proteomic associates of infection mortality in UK Biobank"

**Supplementary Data 1: Genome-wide association study extended methods and quality control data**

**Genotyping and quality control**

UKB contains genomic data for 488,000 participants. Genotype calling was performed on the UK BiLEVE Axiom array (~50,000) and UK Biobank Axiom array (~450,000). There are 805,426 markers in the released genotype data and it was not possible to assay genotypes for ~3% due to insufficient DNA extraction from blood samples. The genotype data was phased using SHAPEIT3 algorithm. Genotype imputation was performed using computational efficient methods combined with the Haplotype Reference Consortium and UK10K haplotype panels. Full details describing the centralised analysis of genetic data has been described elsewhere^1^.

In this study we restricted analyses to autosomal variants and excluded variants with an imputation INFO score <0.3. We filtered out variants with a mean allele frequency (MAF) <0.1%, minor allele count <20 and variant genotyping rate <10%.

We excluded participants with sex discrepancies between reported sex and genetic sex, sex chromosome aneuploidy, outliers for heterozygosity or missing rate and individuals with a missing genotype rate >10%.

We restricted samples to genetically White British individuals using UKB field 22006.

**Genome wide association study**

We conducted a genome-wide association analysis on infection death using regenie, which implements a fast and accurate approach accounting for relatedness and other covariates through a compressed mixed model regression framework^2^. It has good type 1 error control in significant case control imbalance showing good performance with case-control ratios as high as 1:660 in UKB^2^.

In the first step, a selected group of genetic variants is employed to construct an all-encompassing regression model across the genome, effectively capturing a significant portion of the phenotype's variance attributed to genetic influences. Subsequently, in the second phase, the expanded set of variants (imputed) undergoes assessment for correlation with the phenotype. This evaluation occurs conditionally, considering the prediction derived from the earlier regression model. The assessment follows a leave-one-chromosome-out (LOCO) methodology, deliberately avoiding contamination from proximal genetic elements.

GWAS was performed on individuals in UKB without plasma proteomic data using regenie v3.1.1 adjusting for age, sex, genotyping array and the first ten principal components.

**Supplemental Figure 1: GWAS Q-Q plot**

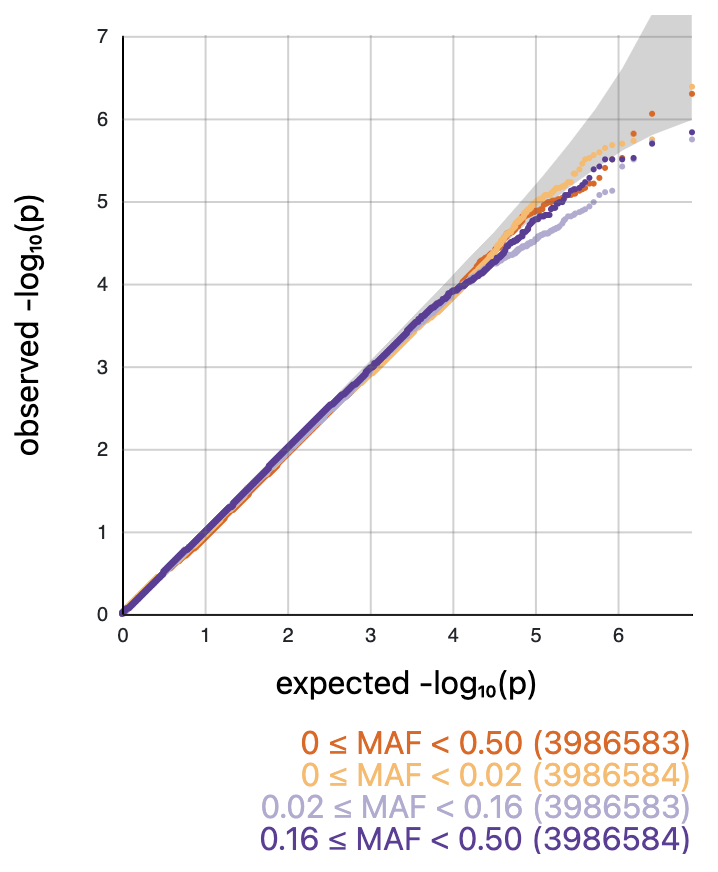

Q-Q plot illustrating distribution of observed versus expected p values in genome-wide association study of infection mortality in UKB individuals without plasma proteomic data

**Supplemental Figure 2: Radial MR**

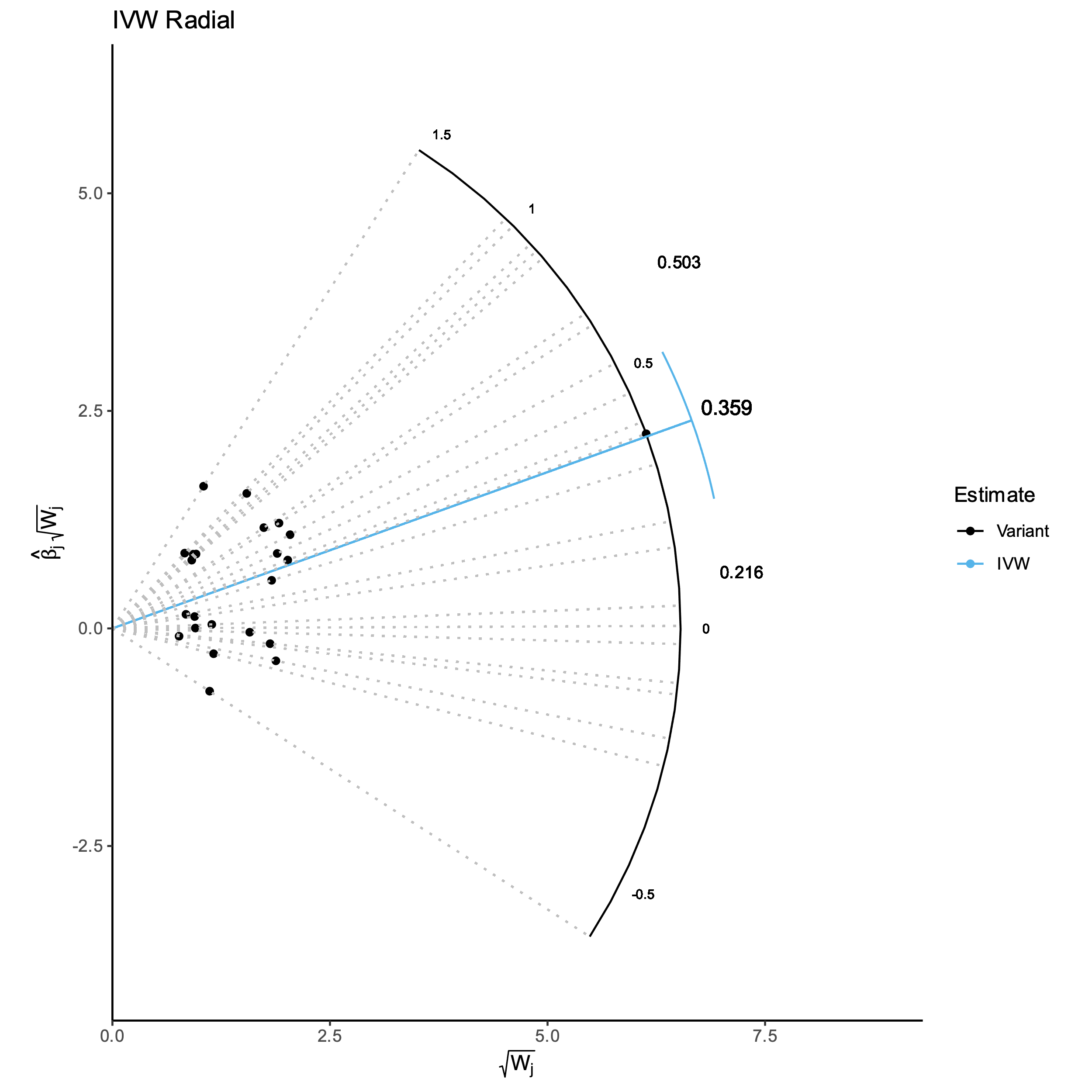

Radial MR plot which shows the MR effect estimate for each SNP on the Y-axis and the weight attached to it on the X-axis. This allows for visualisation of all SNP MR effect estimates, and the identification of outliers which are outside the range of the plot. In this plot we focus on the IVW estimate for the effect of MERTK on infection mortality.
